# Transmission of SARS-CoV-2 Considering Shared Chairs in Outpatient Dialysis: A Real-World Case-Control Study

**DOI:** 10.1101/2021.02.20.21251855

**Authors:** Ravi Thadhani, Joanna Willetts, Catherine Wang, John Larkin, Hanjie Zhang, Lemuel Rivera Fuentes, Len Usvyat, Kathleen Belmonte, Yuedong Wang, Robert Kossmann, Jeffrey Hymes, Peter Kotanko, Franklin Maddux

**Affiliations:** Partners HealthCare, Boston, MA, United States; Fresenius Medical Care, Global Medical Office, Waltham, United States; University of California-Santa Barbara, Santa Barbara, California, United States; Renal Research Institute, New York, United States; Fresenius Kidney Care, Waltham, United States; Fresenius Medical Care North America, Medical Office, Waltham, United States; Icahn School of Medicine at Mount Sinai, New York, United States; Fresenius Medical Care AG & Co. KGaA, Global Medical Office, Bad Homburg, Germany

## Abstract

**Background:** SARS-CoV-2 is primarily transmitted through aerosolized droplets; however, the virus can remain transiently viable on surfaces.

**Objective:** We examined transmission within hemodialysis facilities, with a specific focus on the possibility of indirect patient-to-patient transmission through shared dialysis chairs.

**Design:** We used real-world data from hemodialysis patients treated between February 1^st^ and June 8^th^, 2020 to perform a case-control study matching each SARS-CoV-2 positive patient (case) to a non-SARS-CoV-2 patient (control) in the same dialysis shift and traced back 14 days to capture possible exposure from chairs sat in by SARS-CoV-2 patients. Cases and controls were matched on age, sex, race, facility, shift date, and treatment count.

**Setting:** 2,600 hemodialysis facilities in the United States.

**Patients:** Adult (age ≥18 years) hemodialysis patients.

**Measurements:** Conditional logistic regression models tested whether chair exposure after a positive patient conferred a higher risk of SARS-CoV-2 infection to the immediate subsequent patient.

**Results:** Among 170,234 hemodialysis patients, 4,782 (2.8%) tested positive for SARS-CoV-2 (mean age 64 years, 44% female). Most facilities (68.5%) had 0 to 1 positive SARS-CoV-2 patient. We matched 2,379 SARS-CoV-2 positive cases to 2,379 non-SARS-CoV-2 controls; 1.30% (95%CI 0.90%, 1.87%) of cases and 1.39% (95%CI 0.97%, 1.97%) of controls were exposed to a chair previously sat in by a shedding SARS-CoV-2 patient. Transmission risk among cases was not significantly different from controls (OR=0.94; 95%CI 0.57 to 1.54; p=0.80). Results remained consistent in adjusted and sensitivity analyses.

**Limitation:** Analysis used real-world data that could contain errors and only considered vertical transmission associated with shared use of dialysis chairs by symptomatic patients.

**Conclusions:** The risk of indirect patient-to-patient transmission of SARS-CoV-2 infection from dialysis chairs appears to be low.

**Primary Funding Source:** Fresenius Medical Care North America; National Institute of Diabetes and Digestive and Kidney Diseases (R01DK130067)

## Introduction

Hemodialysis is a lifesaving therapy for individuals with end-stage kidney disease (ESKD). Most ESKD patients are treated in an outpatient setting and require thrice weekly maintenance hemodialysis. Unlike other medical procedures that were canceled or postponed during the initial surge of COVID-19 cases, hemodialysis treatments continued during this period and will continue throughout any subsequent outbreak.(1-3) The pandemic has highlighted several population-specific vulnerabilities to increased mortality risk, including older age, non-white race, and presence of underlying conditions such as diabetes, obesity, cardiovascular disease, and ESKD requiring maintenance hemodialysis.(4-7) Because individuals with ESKD are known to have immunologic deficiencies(8) and higher frequencies of the very same risk factors that predispose humans to adverse outcomes following SARS-CoV-2 infection, their risk for adverse outcomes is compounded.(9-12)

During the pandemic start, outpatient dialysis facilities worldwide implemented enhanced infection control measures to limit transmission of SARS-CoV-2.(13-15) In the United States, mitigation strategies adopted by a national dialysis network included screening before and universal masking while in facility, testing of patients/staff with flu-like symptoms, as well as designating isolation shifts/facilities.(16) While the primary mode of person-to-person transmission of the SARS-CoV-2 virus is through aerosolized droplets,(17, 18) the virus can deposit and remain transiently viable on surfaces, and surface-to-individual transmission represents yet another opportunity for infection.(19-21)

In the United States, outpatient hemodialysis facilities usually accommodate up to 40 chairs/stations per site. Hemodialysis sessions occur in shifts, and each dialysis station (dialysis machine and corresponding chair) in each facility generally accommodates approximately four successive patients each day with dialysis station surfaces cleaned between treatments. Given SARS-CoV-2 may be transmitted while infected individuals remain asymptomatic for several days,(22) we sought to examine transmission dynamics within facilities, with a specific focus on possible indirect transmission through shared dialysis chairs while infected patients were still asymptomatic (i.e. before symptoms or testing SARS-CoV-2 positive). Our primary hypothesis was that chair exposure does not confer a higher risk to the patient sitting on the same chair immediately after an infected patient.

## Methods

### Participants

We used data from all adult (age ≥18 years) in-center hemodialysis patients treated at the national network of dialysis clinics (Fresenius Kidney Care (FKC), Waltham, MA, United States) between February 1, 2020 and June 8, 2020. Patient data were de-identified for analysis, which was performed under a regulatory protocol reviewed by New England Institutional Review Board (Needham Heights, MA, United States; NEIRB WO#: 17-1349084-1); the analysis was approved under Exempt Category (45 CFR 46.104-d4ii) and consent was not required. Analysis was conducted in adherence with Declaration of Helsinki.

### Clinic SARS-CoV-2 Screening and Testing Practices

The national dialysis provider identified the first patients (n=2) with SARS-CoV-2 on March 3, 2020. Since late February 2020, the provider universally adopted modified mitigation measures in response to the pandemic, which included documented screening prior to admittance for patients, staff, and people entering the facility. Patients were provided with and required to wear a surgical face mask during screening and while in facility. Dialysis staff were required to wear surgical face masks, face shields, gowns, and gloves during direct patient care.

Trained healthcare screeners asked standardized questions regarding flu-like symptoms (e.g., fever, respiratory symptoms), recent travel, close-contact to people with COVID-19. Patients/staff without signs/symptoms were admitted. Any patients/staff who presented with signs/symptoms of a flu-like illness or had a body temperature ≥37.8°C failed screening.

Symptomatic patients had their treatments moved to isolation shifts dedicated to persons under investigation (PUI) where staff universally performed nasopharyngeal swabs for SARS-CoV-2 RT-PCR testing. Any patients who failed screening due to recent exposure to someone with COVID-19 were moved into unique isolation shifts apart from symptomatic PUI shifts, if possible. If exposed patients presented with flu-like symptoms while under 14 days of isolation, they were moved to a PUI isolation shift and received RT-PCR testing. PUIs testing SARS-CoV-2 positive were moved to dedicated COVID-19 shifts. PUI and SARS-CoV-2 positive patients were required to have two negative RT-PCR tests >24 hours apart to return to the general hemodialysis population. Isolation facilities or shifts were designated based on the number of suspected/confirmed SARS-CoV-2 patients in geographies. Similar processes were implemented for facility staff; symptomatic/exposed staff were required to quarantine at home, symptomatic staff received RT-PCR testing and were required to have negative test results or be cleared by their healthcare provider before returning to work.

### Facility Infection Control Procedures

Per the provider’s standard disinfection protocol, chairs and outside surfaces of the hemodialysis machine are cleaned between patients using 1:100 bleach; this applies to general and isolation shifts. Internal components of dialysis machines are cleaned according to company policy and CDC guidelines(23), which includes daily acid (i.e. vinegar) and heat disinfection and weekly bleach disinfection. Cleaning procedures take place between successive treatments (average of 90±68 minutes between patients in study period). Patient facing rooms (e.g., lobby, restroom) are thoroughly cleaned during dialysis shift changes.

### Dialysis Station Designation

The machine and station number used for each hemodialysis treatment was documented in electronic medical record. Station number indicates the physical location in the facility where treatment occurs. Chairs are not numbered, however, they are generally not moved from one physical station to another other than: First, if a patient weighs >350 pounds (>159 kg), a special chair is allotted; Second, if a patient had a bleeding episode, a chair may be moved for enhanced cleaning; Third, if a chair breaks during a treatment, a different chair will be assigned; Finally, prior to SARS-CoV-2 outbreak, floors were periodically waxed, hence chairs may have been moved. Examining frequencies of other scenarios that would ‘disconnect’ the chair from the dialysis station yielded insignificant occurrences. Therefore, station number was used as a proxy for chair number with several sensitivity analyses conducted to confirm findings.

### Statistical Analyses

Our primary analysis used a retrospective case-control design,(24) where each SARS-CoV-2 patient (case) was matched with a non-SARS-CoV-2 patient (control) in the same shift. The exposure was defined as hemodialysis in a chair immediately after a SARS-CoV-2 positive patient sat in the same chair during a ‘shedding period’. For a matched case-control pair, the ‘shedding period’ was defined as −3 days before to +7 days after the earliest recorded SARS-CoV-2 presentation or test date; this was the estimated timeframe in which a positive patient or ‘donor’ may have spread SARS-CoV-2 to the next patient that sat in the chair.(25, 26) The ‘shedding period’ window was chosen considering some reports were missing symptom date (only SARS-CoV-2 positive test date), as well as the possibility of a recall bias in recorded symptom date. We performed sensitivity analyses around this window to reduce the likelihood this assumption influenced results. In all analyses we confirmed the ‘donor’ patient was treated in the chair and was not yet transferred to an isolation shift/facility.

The exposure period for a non-SARS-CoV-2 ‘recipient’ patient was defined as −14 days to −1 day before each ‘donor’ patient’s ‘shedding period’; this was the estimated timeframe in which a ‘recipient’ patient could have contracted SARS-CoV-2 from sitting in a chair after a ‘donor’ patient.(27) In all analyses, we excluded data from isolation shifts/facilities, as well as treatments from the first SARS-CoV-2 patient(s) in each facility who would not have had a known opportunity to contract the virus from exposure at the facility. Furthermore, facilities with <200 treatments during the observation period were excluded as it indicated the facility was newly opened or closed, precluding a thorough analysis of potential for transmission.

For each SARS-CoV-2 positive patient (case), we identified their most recent dialysis treatment during their exposure period, identified non-SARS-CoV-2 patients who were treated in that same facility on that day, and then selected a control as the patient who has the best match with the case based on shift time, number of treatments during exposure period, gender, age, and race, in that hierarchical order.

We fitted four conditional logistic regression models: Model 1: exposure status only; Model 2: in addition to Model 1, age, dialysis vintage, sex, race,; Model 3: in addition to Model 2, body mass index (BMI), central venous catheter (CVC) dialysis access, diabetes, chronic obstructive pulmonary disease (COPD); Model 4: in addition to Model 3, county-level 2018 Five-Year Median Household Income and Gini Coefficient (a ratio of household inequality ranging from 0 (complete income equality) to 1 (complete income inequality).(28-30) Analyses were performed using R 3.6.0 (R Foundation for Statistical Computing, Vienna, Austria). The R package survival was used to fit conditional logistic regression models.

We performed sensitivity analyses that varied the definitions of ‘shedding period’ and ‘exposure period.’ The ‘shedding period’ was modified to −3 days before to +3 days after the recorded SARS-CoV-2 date and to −5 before to +10 days after the recorded SARS-CoV-2 date. The exposure period was modified to −14 days before to −3 days before the recorded SARS-CoV-2 date.

As a secondary analysis, we performed a prospective analysis. For each facility we identified the first chair occupied by a shedding SARS-CoV-2 patient and labeled other chairs in the same shift of the same facility on the same day as non-SARS-CoV-2 chairs. We then identified patients who sat in SARS-CoV-2 and non-SARS-CoV-2 chairs during the next shift and recorded the proportion of patients who subsequently developed SARS-CoV-2 within 14 days. Finally, we tested whether there was a difference in the proportion of patients with confirmed SARS-CoV-2 in the next 14 days between those who occupied SARS-CoV-2 chairs compared to non-SARS-CoV-2 chairs.

## Results

We considered data from 170,234 adult hemodialysis patients treated at 2,600 outpatient facilities for matching and identification of case-control pairs (**Figure 1**). Among 636,758 total dialysis shifts examined, 8,948 (1.41%) shifts had at least one SARS-CoV-2 patient. Percent of facilities with 0, 1, 2, and ≥3 SARS-CoV-2 positive patients was 48.3%, 20.2%, 10.0%, and 21.5%, respectively (**Figure 2**). SARS-CoV-2 patients were on average 64 years old, 44% were female, and undergoing hemodialysis for approximately 4 years (**Table 1**). There was a higher proportion of SARS-CoV-2 patients with a Black race and diabetes. Hospitalizations occurred in 72.5% (3466/4782) of SARS-CoV-2 positive patients compared to 28.3% (46,796/165,452) of those who tested negative, or never had flu-like symptoms and were not tested.

**Figure 1:**
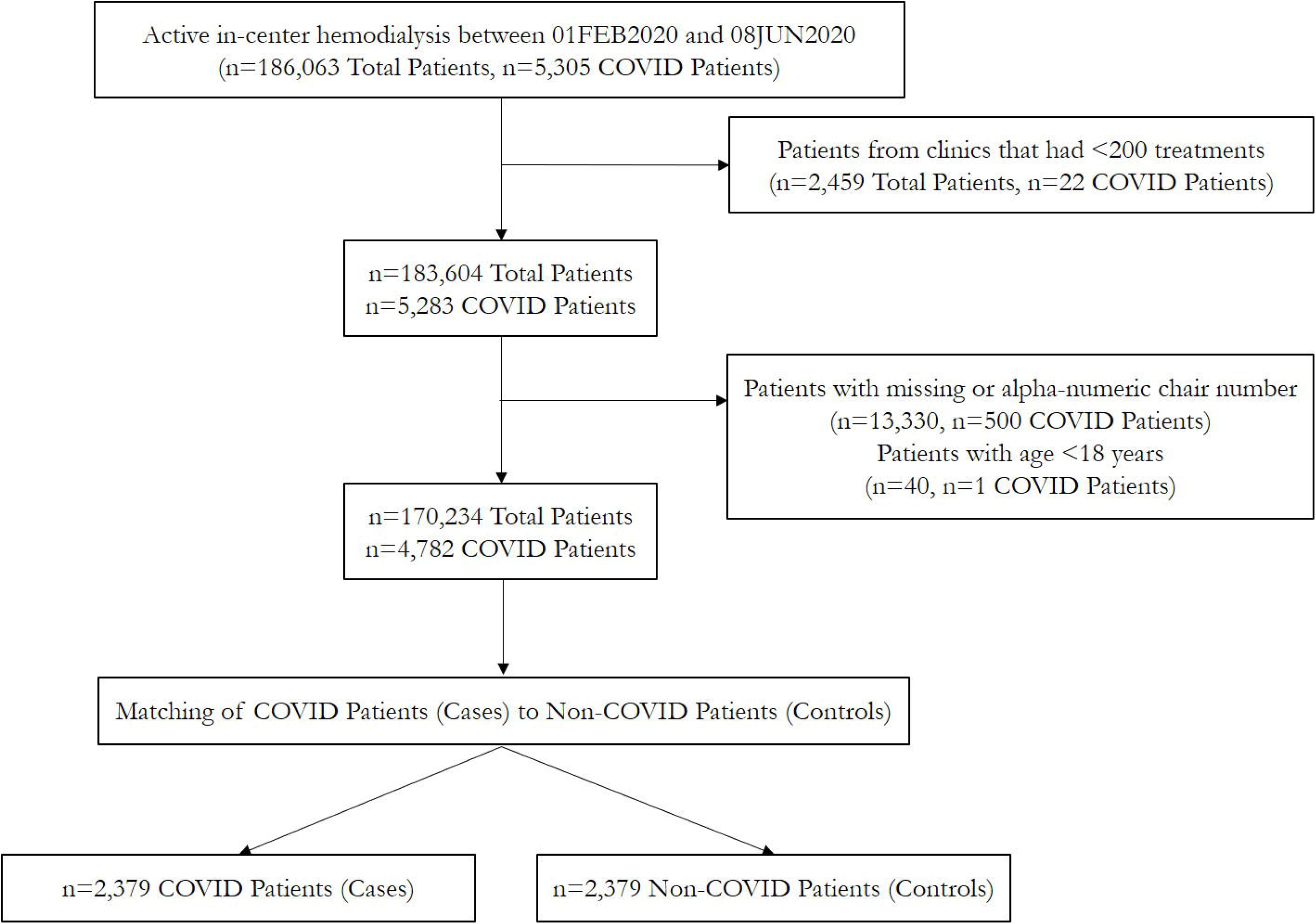
Patient flow diagram

**Figure 2:**
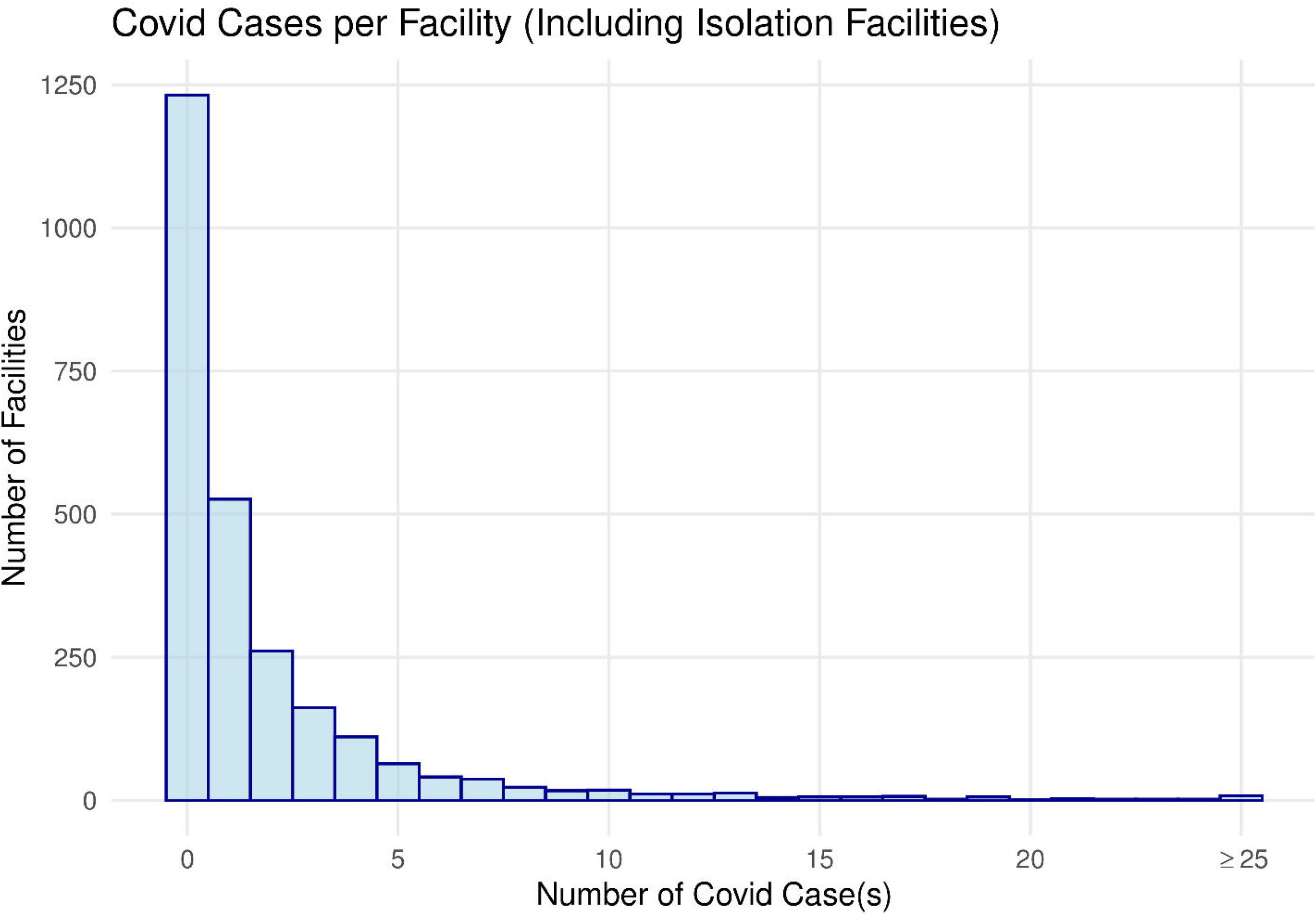
Histogram of number of COVID-19 cases per facility. ^a^Clinics with <200 treatments were excluded as it indicates newly opened or closed clinic; a clinic with one patient would have ∼50 dialysis treatments during this period. ^b^Given that station number is a manually entered field, we chose to exclude clinics that had alpha-numeric station locations that represent entry errors since numeric station numbers are considered standard practice.

**Table 1.** Hemodialysis patient characteristics overall and after case-control matching by COVID-19 status during study period.

|  | Full Patient Population |  |  |  | Matched Case and Control Patients |  |
| --- | --- | --- | --- | --- | --- | --- |
|  | All Patients | COVID Negative | COVID Positive | p-value<sup>a,b</sup> | COVID Negative Controls | COVID Positive Cases |
| **Total Patients** | 170,234 | 165,452 | 4,782 |  | **2,379** | **2,379** |
| Mean Age at study start (yr, sd)<sup>a</sup> | 63.5 (14.2) | 63.5 (14.2) | 64.3 (13.8) | 0.0002 | 63.7 (12.9) | 64.2 (13.8) |
| Mean Time on Dialysis at study start (yr, sd)<sup>a</sup> | 4.2 (4.1) | 4.2 (4.1) | 4.3 (4.0) | 0.0569 | 4.2 (4.1) | 4.4 (4.1) |
| Sex<sup>b</sup> |  |  |  |  |  |  |
| Male (%) | 97,688 (57) | 94,991 (57) | 2,697 (56) | 0.1633 | 1,334 (56) | 1,345 (57) |
| Female (%) | 72,546 (43) | 70,461 (43) | 2,085 (44) | 0.1633 | 1,045 (44) | 1,034 (43) |
| Race<sup>b</sup> |  |  |  |  |  |  |
| White (%) | 72,578 (43) | 70,736 (43) | 1,842 (39) | <0.0001 | 888 (37) | 888 (37) |
| Black (%) | 46,060 (27) | 44,455 (27) | 1,605 (34) | <0.0001 | 848 (36) | 833 (35) |
| Other (%) | 6,720 (4) | 6,579 (4) | 141 (3) | 0.0003 | 75 (3) | 70 (3) |
| Unknown (%) | 44,876 (26) | 43,682 (26) | 1,194 (25) | 0.0266 | 568 (24) | 588 (25) |
| BMI (kg/m<sup>2</sup>, sd)<sup>a</sup> | 29.9 (7.7) | 29.9 (7.7) | 29.9 (7.9) | 0.7699 | 29.1 (7.1) | 29.8 (7.9) |
| CVC used during study (%)<sup>b</sup> | 30,077 (18) | 29,112 (17) | 965 (20) | <0.0001 | 395 (17) | 450 (19) |
| Diabetes (%)<sup>b</sup> | 70,745 (42) | 68,525 (41) | 2,220 (46) | <0.0001 | 964 (41) | 1,119 (47) |
| COPD Diagnosis (%)<sup>b</sup> | 16,331 (10) | 15,874 (10) | 457 (10) | 0.9305 | 202 (8) | 211 (9) |
| Mean Number of Treatments Per Patient during study period (sd)<sup>a</sup> | 48 (13) | 48 (13) | 42 (12) | <0.0001 | 49 (8) | 43 (11) |
| Five Year Median HH Income (sd)<sup>a</sup> | \$58,702 (16,596) | \$58,530 (16,519) | \$64,627 (18,102) | <0.0001 | \$65,563 (18,159) | \$65,534 (18,416) |
| Five Year Gini Coefficient (sd)<sup>a</sup> | 0.47 (0.03) | 0.47 (0.03) | 0.48 (0.04) | <0.0001 | 0.48 (0.04) | 0.48 (0.04) |

<sup>a</sup> For continuous variables, p-values result from two-sided t-test $H_0$ : mean values for COVID negative patients are equal to mean values for COVID positive patients in the full population.
<sup>b</sup> For categorical variables, p-values result from Chi-Square test $H_0$ : proportion in COVID negative patients is equal to proportion in COVID positive patients. Body mass index (BMI); central venous catheter (CVC); chronic obstructive pulmonary disease (COPD); household (HH); year (yr); standard deviation (sd).

We identified matches between 2,379 cases and 2,379 controls in 753 facilities (**Table 1**). Among matched cases-control pairs, 1.30% (95%CI 0.90%, 1.87%) of cases and 1.39% (95%CI 0.97%, 1.97%) of controls had exposure to a chair previously sat in by a shedding SARS-CoV-2 patient. This risk of exposure to a SARS-CoV-2 chair was not statistically different in cases compared to controls (OR=0.94, 95%CI 0.57, 1.54, p=0.80), and results remained consistent after adjusting for potential confounding factors (**Table 2**).

**Table 2.**
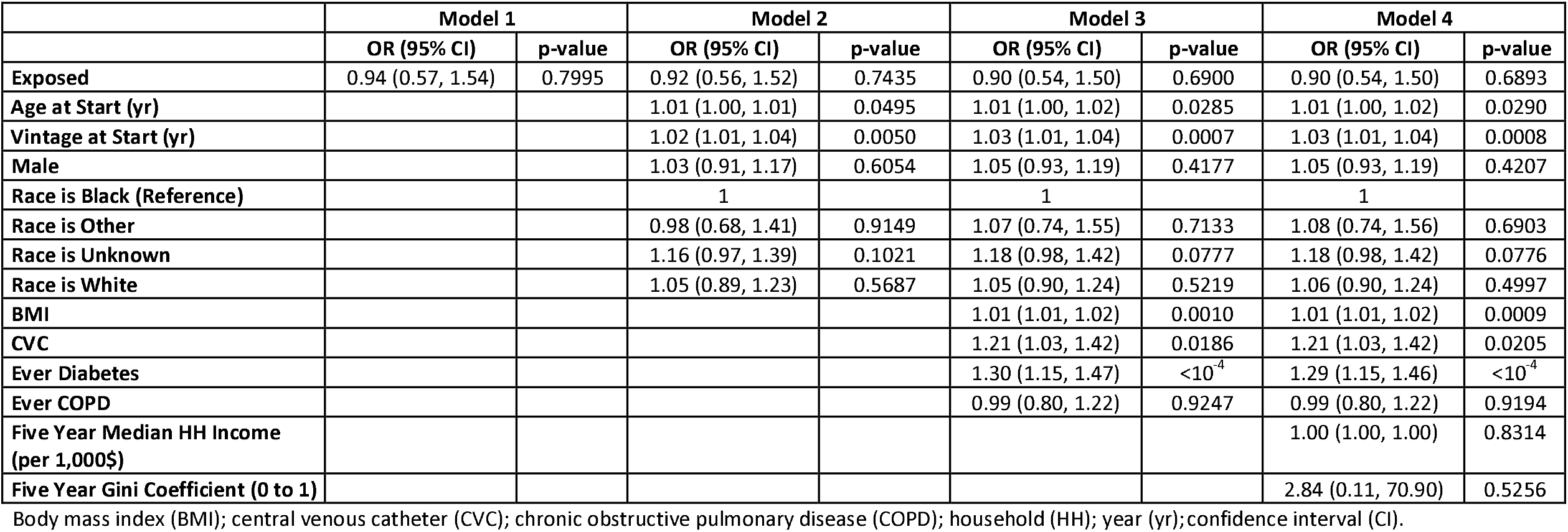
Four conditional logistic regression model results.

|  | Model 1 |  | Model 2 |  | Model 3 |  | Model 4 |  |
| --- | --- | --- | --- | --- | --- | --- | --- | --- |
|  | OR (95% CI) | p-value | OR (95% CI) | p-value | OR (95% CI) | p-value | OR (95% CI) | p-value |
| Exposed | 0.94 (0.57, 1.54) | 0.7995 | 0.92 (0.56, 1.52) | 0.7435 | 0.90 (0.54, 1.50) | 0.6900 | 0.90 (0.54, 1.50) | 0.6893 |
| Age at Start (yr) |  |  | 1.01 (1.00, 1.01) | 0.0495 | 1.01 (1.00, 1.02) | 0.0285 | 1.01 (1.00, 1.02) | 0.0290 |
| Vintage at Start (yr) |  |  | 1.02 (1.01, 1.04) | 0.0050 | 1.03 (1.01, 1.04) | 0.0007 | 1.03 (1.01, 1.04) | 0.0008 |
| Male |  |  | 1.03 (0.91, 1.17) | 0.6054 | 1.05 (0.93, 1.19) | 0.4177 | 1.05 (0.93, 1.19) | 0.4207 |
| Race is Black (Reference) |  |  | 1 |  | 1 |  | 1 |  |
| Race is Other |  |  | 0.98 (0.68, 1.41) | 0.9149 | 1.07 (0.74, 1.55) | 0.7133 | 1.08 (0.74, 1.56) | 0.6903 |
| Race is Unknown |  |  | 1.16 (0.97, 1.39) | 0.1021 | 1.18 (0.98, 1.42) | 0.0777 | 1.18 (0.98, 1.42) | 0.0776 |
| Race is White |  |  | 1.05 (0.89, 1.23) | 0.5687 | 1.05 (0.90, 1.24) | 0.5219 | 1.06 (0.90, 1.24) | 0.4997 |
| BMI |  |  |  |  | 1.01 (1.01, 1.02) | 0.0010 | 1.01 (1.01, 1.02) | 0.0009 |
| CVC |  |  |  |  | 1.21 (1.03, 1.42) | 0.0186 | 1.21 (1.03, 1.42) | 0.0205 |
| Ever Diabetes |  |  |  |  | 1.30 (1.15, 1.47) | <10<sup>-4</sup> | 1.29 (1.15, 1.46) | <10<sup>-4</sup> |
| Ever COPD |  |  |  |  | 0.99 (0.80, 1.22) | 0.9247 | 0.99 (0.80, 1.22) | 0.9194 |
| Five Year Median HH Income (per 1,000\$) | | | | | | | 1.00 (1.00, 1.00) | 0.8314 |
| Five Year Gini Coefficient (0 to 1) |  |  |  |  |  |  | 2.84 (0.11, 70.90) | 0.5256 |

Body mass index (BMI); central venous catheter (CVC); chronic obstructive pulmonary disease (COPD); household (HH); year (yr); confidence interval (CI).

We then performed a series of sensitivity analyses, first modifying the ‘shedding period’ and then modifying the ‘exposure period.’ Modifying the ‘shedding period’ of the donor patient to −3 days before to +3 days after the recorded SARS-CoV-2 date, we identified 2,370 matched case-control pairs; 1.22% (95%CI 0.84%, 1.78%) of cases and 1.22% (95%CI 0.84%, 1.78%) of controls had exposure to a chair sat by a shedding SARS-CoV-2 patient (estimates were identical). Exposure risk between cases and controls was not significant (OR=1.00, 95%CI 0.59, 1.69, p=1.00). Modifying the ‘shedding period’ of the donor patient to −5 days before to +10 days after the recorded SARS-CoV-2 date, we identified 2,516 matched case-control pairs, 2.34% (95%CI 1.80%, 3.04%) of cases and 2.38% (95%CI 1.84%, 3.08%) of controls had exposure to a chair sat on by a shedding SARS-CoV-2 patient. Exposure risk did not differ between cases and controls (OR=0.98, 95%CI 0.68, 1.42, p=0.92). Finally, modifying the ‘recipients’ ‘exposure period’ from −14 to −3 days before the recorded SARS-CoV-2 date, we identified 2,335 matched case-control pairs; 1.24% (95%CI 0.85%, 1.80%) of cases and 1.28% (95%CI 0.88%, 1.85%) of controls had exposure to a chair sat in by a shedding SARS-CoV-2 patient. Exposure risks between cases and controls was not different (OR=0.97, 95% CI 0.57, 1.62, p=0.89). No results of sensitivity analyses materially changed when adjusted for various confounders.

Finally, we examined exposure risk prospectively. Among 343 and 4552 patients who sat in the next shift in a SARS-CoV-2 chair and a non-SARS-CoV-2 chair, respectively, and 2 (0.58%) and 42 (0.92%) contracted SARS-CoV-2 in the following 14 days (χ^2^ =0.1197, p=0.73).

## Discussion

Transmission of SARS-CoV-2 attached to inanimate surfaces is a justified concern since the virus can remain viable on surfaces.(19) Routine capture of real-world data at a national network of outpatient hemodialysis facilities afforded a unique opportunity for examination of transmission between patients in a closed medical environment. Hemodialysis patients sit for approximately 3-5 hours in chairs during thrice weekly sessions. While strict and highly standardized cleaning procedures are in place, there are concerns about possible transmission of SARS-CoV-2 from an infected patient to the dialysis station (chair and/or machine) and from there to the next patient. Our analysis assessed SARS-CoV-2 transmission risk in patients dialyzed at the same station immediately after an infected patient. Our results indicate dialysis chairs and their corresponding machines are unlikely intermediate vectors of patient-to-patient transmission.

SARS-CoV-2 can remain viable on inanimate surfaces depending on temperature, humidity, and material. On polypropylene plastics and stainless steel, the virus can be viable for up to 72 hours.(19) On hard surfaces, disinfection with at least 70% ethanol, 0.5% hydrogen peroxide, or 0.0525% sodium hypochlorite (1:100 bleach) was shown to inactivate ≥99% of SARS-CoV-2 in most cases and at least inactivate >90% of the virus.(31) Dialysis chair, arms, and all patient contact surfaces are covered by Medical Grade Vinyl (polyvinyl chloride-PVC) designed to withstand most disinfectants. Meticulous cleaning of chairs with 1:100 bleach between patients may have conferred protection to subsequent sitters. Although not tested specifically, it is possible our findings are applicable to potential transfer surfaces such as chairs in medical/dental offices, waiting rooms, airports, theaters, transports, and places of worship provided cleaning measures mimic those used in dialysis facilities.

Our study has several strengths including use of a comprehensive data set with documentation of SARS-CoV-2 status and records of standard parameters of dialysis shifts, machines, and stations. A series of sensitivity analyses and a prospective analysis showed consistency across analyses. Overall, most facilities had 0 to 1 confirmed infected patient, which suggests within-center transmission of SARS-CoV-2 is low.

Our study has some limitations. We had no data to assess potential transmission in shared vehicle transport to facility. Since most facilities had at only 0 or 1 infected patient, and since patients scheduled in similar shifts are usually transported together in shared personal or medical/public transport vehicles, infection during transport is not anticipated to have affected our results. Second, our analysis considers only symptomatic patients. Therefore, we cannot exclude the possibility of transmission from asymptomatic carriers who never developed symptoms that warranted RT-PCR testing. While asymptomatic spread is possible, individuals who remain asymptomatic throughout their entire course of SARS-CoV-2 infection are typically >30 years younger than the average dialysis patient, who is usually in the mid-sixties, and the secondary attack rate among asymptomatic carriers is generally lower than symptomatic carriers.(32-34) Finally, while we believe the fidelity of linking a specific dialysis machine to a specific chair was maintained, and thus we could examine vertical transmission, we could not examine the potential for horizontal transmission during a single shift because the proximity of one station to another could not be ascertained by the available data. That said, stations are spaced apart, ideally 6 feet, and patients were required to wear masks, making the likelihood of such transmission low.

We also acknowledge that in these challenging times, occasional erroneous documentation of station numbers linked to dialysis machines may have occurred. However, based on our findings, we consider vertical spread of SARS-CoV-2 in hemodialysis facilities unlikely, and that like studies examining transmission among hospital workers, household and restaurant contacts represent a more important site of spread.(35) Lastly, our data do not allow us to assess the possibility of vertical transfer between shift with staff members serving as vectors.

In summary, in many dialysis facilities across the United States that continued to deliver lifesaving chronic hemodialysis treatments in the pandemic, our findings suggest SARS-CoV-2 transmission risk is low for patients occupying the same chair as someone likely shedding the virus. These results may have been due to the rigorous cleaning protocols that take place in between patients.

## Data Availability

This analysis was conducted on a deidentified dataset created by Fresenius Medical Care North America. This dataset is restricted to use under a formal collaboration agreement between Fresenius Medical Care North America and the investigators' institutions. The deidentified dataset is not publicly available.

## Acknowledgments

We would like to thank Vladimir M Rigodon for assistance with regulatory protocol composition.

